# Support needs and barriers to accessing support: Baseline results of a mixed-methods national survey of people bereaved during the COVID-19 pandemic

**DOI:** 10.1101/2021.06.11.21258575

**Authors:** E. Harrop, S. Goss, D. Farnell, M. Longo, A. Byrne, K. Barawi, A. Torrens-Burton, A. Nelson, K. Seddon, L. Machin, E. Sutton, A. Roulston, A. Finucane, A. Penny, K.V. Smith, S. Sivell, L.E. Selman

**Affiliations:** Cardiff University, Marie Curie Research Centre, Division of Population Medicine, Cardiff, UK; Cardiff University, School of Dentistry, Cardiff, UK; Cardiff University, PRIME Centre, Division of Population Medicine, Cardiff, UK; Keele University, Keele, UK; University of Bristol, Palliative and End of Life Care Research Group, Population Health Sciences, Bristol Medical School, Bristol, UK; Queen’s University, Belfast, Northern Ireland; Clinical Psychology, School of Health in Social Science, University of Edinburgh, UK; National Bereavement Alliance/Childhood Bereavement Network, UK; Centre for Anxiety Disorders and Trauma, Department of Experimental Psychology, University of Oxford, UK

**Author notes:** Corresponding author: Dr Emily Harrop, Marie Curie Palliative Care Research Centre, Cardiff University, 8^th^ Floor Neuadd Meirionydd, Heath Park Way, Cardiff CF14 4YS.

**Keywords:** Bereavement, Grief, Pandemics, Coronavirus Infections, Social Support, Bereavement Services

## Abstract

**Background:** The COVID-19 pandemic is a mass bereavement event which has profoundly disrupted grief experiences. Understanding support needs and access to support among people bereaved at this time is crucial to ensuring appropriate bereavement support infrastructure.

**Aim:** To investigate grief experiences, support needs and use of formal and informal bereavement support among people bereaved during the pandemic.

**Design:** Baseline results from a longitudinal survey. Support needs and experiences of accessing support are reported using descriptive statistics and thematic analysis of free-text data.

**Setting/Participants:** 711 adults bereaved in the UK between March-December 2020, recruited via media, social media, national associations and community/charitable organisations.

**Results:** High-level needs for emotional support were identified. Most participants had not sought support from bereavement services (59%, n=422) or their GP (60%, n=428). Of participants who had sought such support, over half experienced difficulties accessing bereavement services (56%, n=149)/GP support (52%, n=135). 51% reported high/severe vulnerability in grief; among these, 74% were not accessing bereavement or mental-health services. Barriers included limited availability, lack of appropriate support, discomfort asking for help, and not knowing how to access services. 39% (n=279) experienced difficulties getting support from family/friends, including relational challenges, little face-to-face contact, and disrupted collective mourning. The perceived uniqueness of pandemic bereavement and wider societal strains exacerbated their isolation.

**Conclusions:** People bereaved during the pandemic have high levels of support needs alongside difficulties accessing support. We recommend increased provision and tailoring of bereavement services, improved information on support options, and social/educational initiatives to bolster informal support and ameliorate isolation.

**Key statements:** *What is already known about the topic?:*

– Features of pandemic bereavement, such as traumatic death experiences, exacerbate family distress and add to the complexity of grief.
– In pre-pandemic times most people mainly relied on the informal support of friends and family to cope with their bereavement, but an estimated 40% required more formal therapeutic support from bereavement or mental health services.
– Bereaved people experience difficulties getting the support that they need from bereavement services and their social networks.

*What this paper adds:*

– Participants had high level needs for emotional support, especially dealing with/expressing feelings, with 51% experiencing high or severe vulnerability in grief; however, 74% of this group were not accessing formal bereavement service or mental health support.
– Most participants had not tried to access bereavement services, for reasons such as lack of appropriate support, discomfort in asking for help and uncertainty of how to access services; of the 41% who tried, 56% experienced difficulties such as long waiting lists or ineligibility.
– A substantial proportion of people (39%) reported difficulties accessing support from friends and family; reduced in-person contact affected the perceived quality of support and disrupted collective mourning practices, whilst the wider social difficulties of the pandemic compounded feelings of isolation.

*Implications for policy and practice:*

– Further investment in the provision of tailored bereavement support is needed to meet the diverse needs and backgrounds of bereaved people, including support that is culturally and crisis/context competent, and group-based support for those with shared experiences and characteristics.
– To raise awareness of support options, information on grief and bereavement services should be provided proactively following a death and made available in online and community settings, with GPs and other primary care providers better resourced to signpost to appropriate support.
– Following compassionate communities approaches, expanded provision of informal community-based support and activities could help with isolation, whilst longer-term educational and societal initiatives are needed to bolster community support for people experiencing death, dying and bereavement.

## Background

The COVID-19 pandemic has resulted in widespread bereavement on an unprecedented global scale. Lack of access to, and physical contact with, loved ones at the time of death, restrictions surrounding funerals and the sudden nature of most Covid-19 deaths have caused high levels of distress to those bereaved during the pandemic(1-4). Traumatic end-of-life and death experiences add to the complexity of grief(5-11), whilst limited access to usual support networks and severe societal disruption are also likely to increase risks of poor bereavement outcomes(11-14).

Bereavement support is a core part of health and social care provision, and is of heightened importance at times of mass bereavement(12,15,16). NICE guidance and public health approaches to bereavement care recognize the differing needs of bereaved people and recommend a tiered approach to support(15,17,18). The first tier includes universal access to information on grief and available support, recognising that (in pre-pandemic times) c.60% of bereaved people cope without formal intervention, supported by existing social networks(15,17,18). The second tier includes structured, reflective support, beneficial for those with moderate needs, estimated at c.30% of the bereaved population(15,17-19). Third tier support, including specialist grief, mental health and psychological interventions, should be targeted at the small minority (c.10%) of people at high risk of prolonged grief disorder(15,17,18). A review of bereavement interventions following mass bereavement events confirmed the value of social networks, psycho-educational approaches, group-based support and specialist psychological support for those with complex needs, alongside early, proactive outreach to bereaved families(16).

However, there is evidence that bereaved people experience problems getting the right support. These include lack of understanding and compassion amongst family and friends, and difficulties expressing their feelings and needs(20-24). The limited evidence on pandemic bereavement suggests these experiences are intensified by the physical isolation brought about by lockdown and social distancing restrictions, as well as a sense of feeling forgotten(25,26). Disparities between the amount of formal support available and the volume of people who need it have been identified before and during the pandemic(27-29). Barriers to support include lack of information and knowledge of how to get support, and discomfort or reluctance to seek help from services(16,27). Limited awareness of available support and a lack of culturally competent services are particular barriers for people from minority ethnic communities(30,31).

This mixed-methods longitudinal study is the first to investigate bereavement support needs and experiences in the UK during COVID-19, adding to the emerging evidence-base on pandemic bereavement(1-3,11). This paper reports baseline survey results. Using quantitative and qualitative free-text data it describes support needs and experiences of accessing formal and informal support, to inform support provision during and beyond the current crisis.

## Methods

### Study design and aim

Baseline results from a longitudinal survey which aims to investigate the grief experiences, support needs and use of bereavement support by people bereaved during the pandemic. The Checklist for Reporting Results of Internet E-Surveys(32) was followed.

### Survey development

An open web survey (Supplementary file 1) was designed by the research team, which includes a public representative (KS), with input from the study advisory group. It was piloted, refined with 16 public representatives with experience of bereavement and tested by the advisory group and colleagues. Survey design was informed by study aims and previous research(19,24,27,33). Non-randomised open and closed questions covered end-of-life and grief experiences, and perceived needs for, access to and experiences of formal and informal bereavement support. Grief was measured using the Adult Attitude to Grief (AAG) Scale(34), which gives an overall index of vulnerability (0-20 = low vulnerability, 21-23 = high vulnerability, and 24-36 = extreme vulnerability). Most free-text data reported here was from question C4: “If relevant, please briefly describe any difficulties you faced getting support from friends, family or bereavement services”.

### Study procedure

The survey was administered via JISC (https://www.onlinesurveys.ac.uk/) and was open from 28th August 2020 to 5th January 2021. It was disseminated to a convenience sample on social and mainstream media and via voluntary sector associations and bereavement support organisations, including those working with ethnic minority communities. Organisations helped disseminate the voluntary (non-incentivised) survey by sharing on social media, web-pages, newsletters, on-line forums and via direct invitations to potential participants (see Supplementary file 2, example advertisement). For ease of access, the survey was posted onto a bespoke study-specific website with a memorable url (covidbereavement.com). Two participants completed the survey in paper format. Summaries of interim survey results (released November 2020) were posted on the website and provided to interested participants.

### Inclusion criteria

aged 18+; family or close friend bereaved since social-distancing requirements were introduced in the UK (16/03/2020); death occurred in the UK; ability to consent. The initial section of the survey requested informed consent and details data protection (see Supplementary file 1). Via contact and demographic information we identified 12 surveys completed in duplicate; the first completed survey was retained for these participants. Two surveys were excluded as only the consent question had been answered.

### Data analysis

Descriptive statistics and frequency tables were used for demographic and categorical response data. Statistical calculations were carried out using SPSS V26. No statistical correction was carried out. Free-text survey responses were analysed using inductive thematic analysis, involving line-by-line coding in NVivo V12 and identification of descriptive and analytical themes(35). A preliminary coding framework was developed based on a sample of survey responses (ES). The framework was revised and applied in an iterative process (EH,ES,SG,KB) involving independent double coding of 10% of the dataset (70 responses) and regular discussion and cross-checking within the study team. 85% of participants (n=606) provided comments related to experiences of accessing support.

## Results

### Sample Characteristics

711 bereaved participants completed the survey (Table 1). Participants represented diverse geographical areas, deprivation indexes and levels of education. 88.6% of participants were female (n=628); the mean age of the bereaved person was 49.5 years old (SD = 12.9; range 18-90). The most common relationship of the deceased to the bereaved was parent (n=395,55.6%), followed by partner/spouse (n=152,21.4%). 72 people (10.1%) had experienced more than one bereavement since 16^th^ March 2020. 33 people (4.7%) self-identified as from a minority ethnic background.

**Table 1:** Characteristics of the bereaved person. (*multiple bereavements recorded by participants explain discrepancies between overall totals in sibling, child and parent groups and their sub-categories).

| Age (Years) | Mean [Median] | SD | Minimum | Maximum |
| --- | --- | --- | --- | --- |
|  | 49.5 [50.0] | 12.9 | 18 | 90 |
| Gender Identity | Male $n$ (%) | Female $n$ (%) | Other $n$ (%) | |
|  | 74 (10.4%) | 628 (88.6%) | 7 (1%) |  |
| Ethnicity | Non-BAME $n$ (%) | BAME $n$ (%) | | |
|  | 676 (95.3%)<br>White British 438<br>White English 111<br>White Welsh 41<br>Northern Irish 22<br>White Scottish 40<br>Any other white 17<br>White Irish 7 | 33 (4.7%)<br>White and Black Caribbean 12<br>White and Asian 5<br>Indian 4<br>Black Caribbean 4<br>Any other mixed background 3<br>Pakistani 1<br>Bangladeshi 1<br>Arab 1<br>White and Black African 1<br>Any other Asian 1 |  |  |
| Highest Qualification | None or GCSEs<br>*n* (%) | A-level or Apprenticeship or ONC<br>*n* (%) | HND or University Degree<br>*n* (%) |  |
|  | 108 (15.3%) | 132 (18.6%) | 468 (66.1%) |  |
| Was death expected? | Yes *n* (%) | No *n* (%) | Don't Know<br>*n* (%) |  |
|  | 113 (16.0%) | 552 (78.0%) | 43 (6.1%) |  |
| Bereavements in previous year? | Yes *n* (%) | No *n* (%) |  |  |
|  | 158 (22.5%) | 543 (77.5%) |  |  |
| Unemployed during pandemic? | Yes *n* (%) | No *n* (%) |  |  |
|  | 55 (7.9%) | 645 (92.1%) |  |  |
| Region | England | Wales | Scotland | Northern Ireland |
|  | 517 (78.5%) | 63 (9.6%) | 53 (8.0%) | 26 (3.9%) |
| Religious Beliefs |  | *N* | Percentage |  |
|  | Buddhism | 8 | 1.2% |  |
|  | Christian | 251 | 36.7% |  |
|  | Hinduism | 3 | 0.4% |  |
|  | Islam | 5 | 0.7% |  |
|  | Judaism | 6 | 0.9% |  |
|  | Sikhism | 2 | 0.3% |  |
|  | Other or agnostic | 107 | 15.7% |  |
|  | No | 301 | 44.1% |  |
| Relationship of the Deceased Person to the Bereaved* |  | *N* | Percentage |  |
|  | Partner (Male / Female) | 152 (129 / 23) | 21.4% (18.14% / 3.23%) |  |
|  | Parent (Father / Mother) | 395 (218/ 197) | 55.6% (30.7% / 27.7%) |  |
|  | Grandparent | 54 | 7.6% |  |
|  | Sibling (Brother / Sister) | 23 (15 / 10) | 3.2% (2.1%, 1.4%) |  |
|  | Child (Son / Daughter) | 15 (12/ 4) | 2.1% (1.7% / 0.6%) |  |
|  | Other family member | 46 | 6.5% |  |
|  | Colleague or friend | 26 | 3.7% |  |

The mean age of the deceased person was 72.2 years old (SD=16.1; range 4 months gestation to 102 years) (Table 2). 43.8% (n=311) died of confirmed/suspected COVID-19, 21.9% (n=156) from cancer, and 16.7% (n=119) from another life-limiting condition. Most died in hospital (n=410; 57.8%). Questionnaires were completed a median of 152 days (5 months) after the death (range 1-279 days).

**Table 2:** Characteristics of the deceased. (LLC = Life-limiting condition e.g heart disease, COPD, dementia; NLLC = Non-life-limiting condition e.g stroke, heart attack, accident, suicide)

| Age (Years) | Mean [Median] | SD | Minimum | Maximum |
| --- | --- | --- | --- | --- |
|  | 72.2 [74.0] | 16.1 | 0.33 | 102 |
| Cause of Death | COVID *n* (%) | Suspected COVID *n* (%) | Non-COVID *n* (%) |  |
|  | 273 (38.5%) | 38 (5.4%) | 399 (56.2%)<br>Cancer 156 (21.9%)<br>Other LLC 118 (16.7%)<br>Non LLC 112 (15.8%)<br>Don't know 12 (3%)<br>Not specified 1 (0.2%) |  |
| Place of Death |  | *N* | Percentage |  |
|  | In hospital | 410 | 57.7% |  |
|  | In their home | 158 | 22.2% |  |
|  | In a hospice | 37 | 5.2% |  |
|  | In a care home | 91 | 12.8% |  |
|  | Other / Do not Know | 13 | 1.8% |  |

### Support needs and access to formal support

Support needs were assessed in 13 domains (Figure 1, Table 3). In six emotional-support domains, between 50 and 60% of participants reported high/fairly high levels of need. The most common were: ‘dealing with my feelings about the way my loved one died’ (60%), ‘expressing my feelings and feeling understood by others’ (53%), and ‘feelings of anxiety and depression (53%). Over half of participants also demonstrated high or severe levels of overall vulnerability in grief, assessed via the AAG Scale (Severe=28%, high=23%, low=48%, Table 4). Despite this, 72% (n=230) of people with high/severe vulnerability and who were more than 6 weeks bereaved (n=318), were not using formal tier 2/3 support (Figure 2). Most participants had not tried to access support from a bereavement service (59 %, n=422,%) or their GP (60%,n=428); just under a half of whom had high or severe vulnerability (45/44% respectively; n=190). Amongst the 267 participants who sought support from bereavement services, 56% (n=149) experienced access difficulties, with similar proportions observed for GP services (52%,n=135/159).

**Table 3:** Support needs ranked by mean level of need

|  | High or fairly high level of support needed | Moderate level of support needed | Little or no support needed | Mean (95% CI) | Median |
| --- | --- | --- | --- | --- | --- |
| Dealing with my feelings about the way my loved one died | 59.8% | 21.5% | 18.7% | 3.71<br>(3.62 to 3.80) | 4 |
| Dealing with my feelings about being without my loved one | 49.9% | 29.3% | 20.8% | 3.48<br>(3.39 to 3.57) | 3 |
| Expressing my feelings and feeling understood by others | 53% | 23.9% | 23% | 3.48<br>(3.38 to 3.57) | 4 |
| Feeling comforted and reassured | 51.8% | 26.7% | 21.6% | 3.46<br>(3.37 to 3.55) | 4 |
| Feelings of anxiety and depression | 52.8% | 21.1% | 26.1% | 3.45<br>(3.35 to 3.55) | 4 |
| Loneliness and social isolation | 52.0% | 19.1% | 29% | 3.36<br>(3.26 to 3.46) | 4 |
| Finding balance between grieving and other areas of life | 45.0% | 27.9% | 27% | 3.29<br>(3.20 to 3.39) | 3 |
| Regaining sense of purpose and meaning in life | 46.7% | 21.6% | 31.7% | 3.26<br>(3.15 to 3.36) | 3 |
| Managing and maintaining my relationships with friends and family | 36.2% | 26.4% | 37.4% | 2.98<br>(2.88 to 3.08) | 3 |
| Participating in work, leisure or other regular activities (e.g. shopping, housework) | 33.8% | 23.9% | 42.1% | 2.87<br>(2.76 to 2.97) | 3 |
| Getting relevant information and advice e.g. legal, financial, available support | 24.3% | 22.3% | 53.3% | 2.51<br>(2.41 to 2.61) | 2 |
| Practical tasks e.g. managing the funeral, registering the death, other paperwork etc. | 23.5% | 21.7% | 54.7% | 2.48<br>(2.38 to 2.58) | 2 |
| Looking after myself/family e.g. getting food, medication, childcare | 15.2% | 22.8% | 62% | 2.25<br>(2.16 to 2.34) | 2 |

Note that to interpret means and medians: no support = 1; little support = 2; moderate support = 3; fairly high support = 4; and high support = 5)

**Table 4:** Descriptive statistics for the AAG questionnaire(34)

| AAG | Subscales/<br>Total score | *n* | % Missing | Mean (95% CI)<br>[Median] | SD |
| --- | --- | --- | --- | --- | --- |
|  | Overwhelmed | 705 | 0.8% | 8.53 (8.31 to 8.72)<br>[9.00] | 2.79 |
|  | Controlled | 700 | 1.5% | 6.61 (6.41 to 6.82)<br>[7.00] | 2.71 |
|  | Reversed<br>Resilience | 701 | 1.4% | 5.28 (5.07 to 5.49)<br>[5.00] | 2.82 |
|  | IOV | 698 | 1.8% | 20.41 (20.06 to 20.77)<br>[21.00] | 4.77 |
|  | IOV Grouped<br>into<br>Categories | Low *n* (%) | High *n* (%) | Severe *n* (%) |  |
|  |  | 338<br>(48.4%) | 163<br>(23.4%) | 197<br>(28.2%) |  |

Note: The AAG has three subscales (overwhelmed, controlled, and reversed resilience). Subscale scores are calculated by summing the scores for each item for each subject, giving a range from 0 to 12 for each subscale, with increasing scores indicating higher levels of feeling overwhelmed, controlled, and reversed resilience (i.e., vulnerability). The overall index of vulnerability (IOV) is calculated by adding the scores for each subscale together, where IOV score 0-20 = low vulnerability, 21-23 = high vulnerability, and 24-36 = extreme vulnerability.

**Figure 1:**
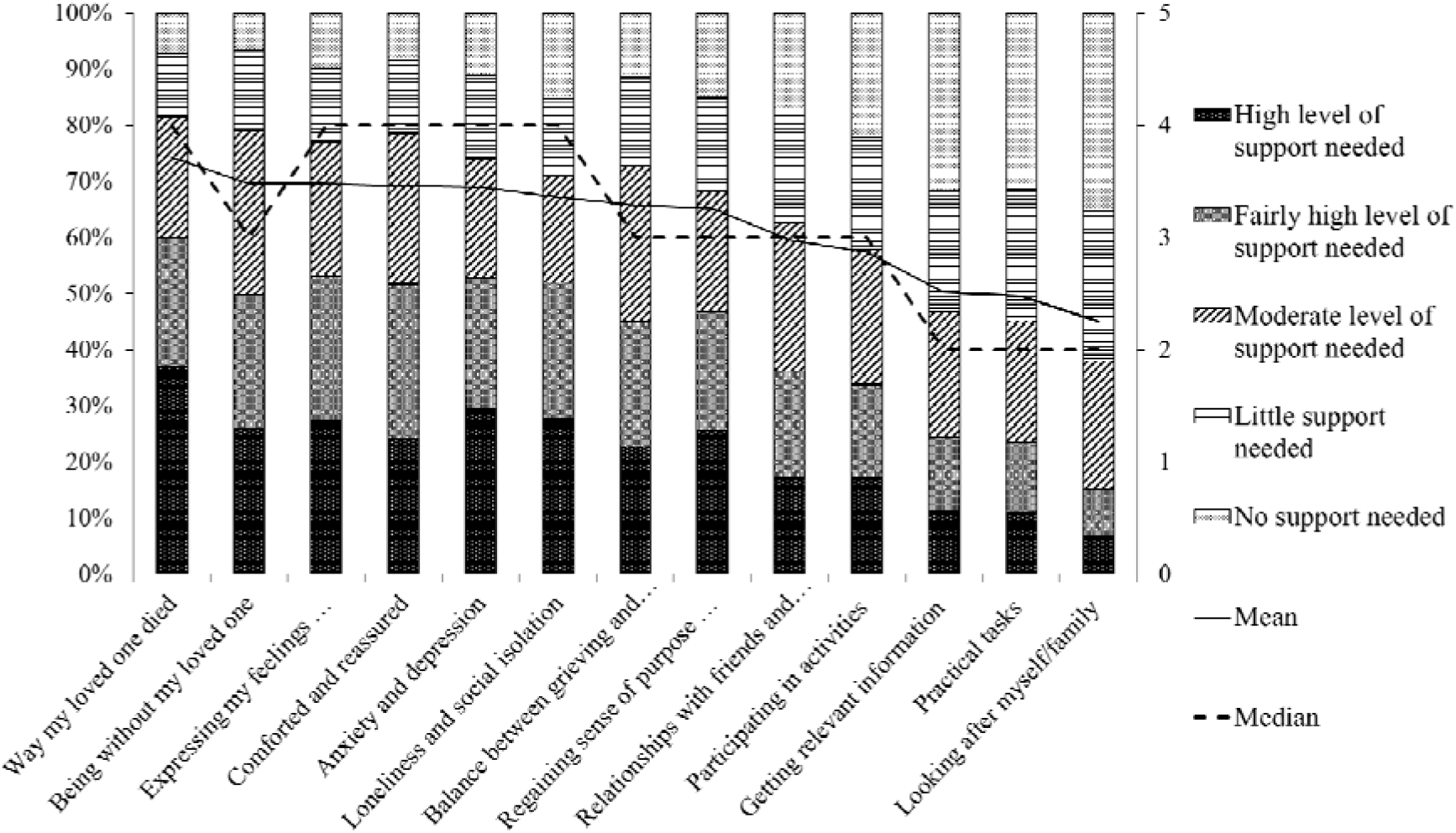
Support needs ranked by mean level of need Note: in order to interpret means and medians, no support = 1; little support = 2; moderate support = 3; fairly high support = 4; and high support = 5

**Figure 2:**
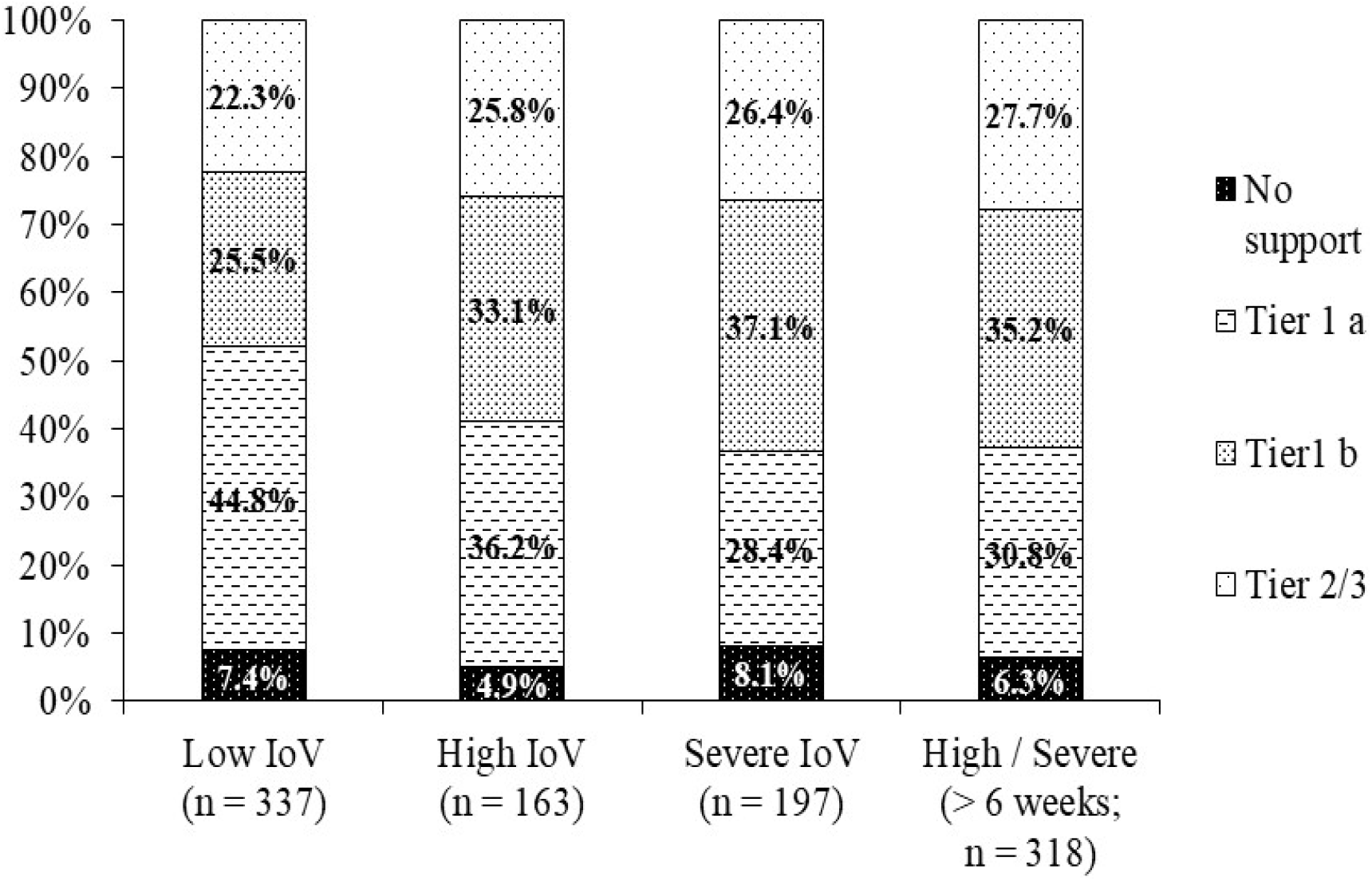
Highest level of support accessed by IOV group using 3 Tiers of Public Health Model (17) Note: Tier 1 a: Friends and family only; Tier 1 b: Informal and information-based support: GP, helpline, online community support, informal support group, other e.g websites, podcasts, self-help material; Tier 2/3: Formal bereavement service and mental health support: One to one support/counselling, bereavements support group/group counselling, mental health support. > 6 weeks; participants who completed questionnaire at least 42 days post-death

Reasons for not using bereavement services included: not needing the support due to adequate support from friends and family (29%,n=207); feeling uncomfortable asking for support (27%, n=195); a perception that the support will not help (18%, n=130); unavailability (15%, n=103), and not knowing how to get support (14%, n=96). Free-text comments expanded upon these reasons, with two main themes identified: availability of (appropriate) support, and knowledge and attitudes towards support use. These themes were represented across gender and ethnic groups; group-specific findings were also identified, described below.

#### Availability of (appropriate) support

Several problems relating to the availability of support were described. Assumptions of over-stretched services dissuaded some participants from trying to get help, whilst others described experiences of unreturned calls, needing to wait a minimum period after the death (6 weeks to 6 months), long waiting lists and post-code-based ineligibility. Some people experienced difficulty getting through to helplines or struggled with discontinuity between call handlers or the limited sessions available. A small minority paid for private counselling to access more timely help.

> *The services are so overloaded that there [are] huge waits to get help. I phoned a local bereavement charity that the hospital recommended when I called them to say that I wasn’t coping well. However, the charity informed me that they would add me to their waiting list, but probably wouldn’t be able to get back to me for 7 months. It made my grief and anxiety even worse knowing that I couldn’t get access to support*. (Bereaved mother, RID696)
>
> *Too long a wait for counselling; don’t feel I have the strength to retell the same story to different people on a help line*. (Bereaved daughter, RID315)

Although many participants who received telephone-or web-based support described positive experiences, some felt uncomfortable discussing sensitive and personal matters remotely. People with children at home or who were working in shared offices reported problems with privacy and having the time and space to access remote support. Some perceived a need for COVID-19 loss-specific support, rather than ‘generic’ support, or reflected on a lack of contextual understanding among support providers. Needs for culturally-relevant support and group-specific support for those with similar experiences were also described; for example widows, young widows, parents, same-sex couples and following particular types of death (COVID-19, childbirth-related, pregnancy loss).

> *I can’t bear the idea of having a bereavement service talk to me about loss in the traditional ways, because losing someone to Covid and in the middle of a lockdown isn’t like other types of loss. What possible advice could they give? There just isn’t any way of observing the sorts of traditions or rituals that would be healing. And how can I move past this when the pandemic is still all around me?* (Bereaved daughter, RID336)
>
> *I wrote to the hospice asking for group support. I definitely NEED to connect with people who have been bereaved during lockdown not particularly because of covid. This hasn’t been possible. The hospice closed their groups and when they were able to offer a zoom group I had returned to work and couldn’t make the time they offered me. I am still waiting for a place in a group at a time I can attend. As I write I wonder if they have forgotten about me. I would also want to meet with others who have lost a same sex partner (not necessarily the same group). Again I can’t find a group that’s meeting*. (Bereaved wife/partner, RID487)

#### Knowledge and attitudes towards support use

Other barriers related to a lack of information about how to get support, feeling too uncomfortable or upset to seek formal support, feeling unable to open up to ‘strangers’ or unsure how bereavement support could help. People who lost elderly parents to long-term illnesses (and in some cases COVID-19), felt less entitled or worthy of support, whilst a woman grieving her female partner explained ‘Because of our relationship I do not know where to turn to for help’ (RID667). Some participants who answered the survey soon after the death considered it too early to be thinking about accessing formal support, but others perceived a need for early intervention.

> *I feel awkward making a phone call to say that I am struggling, especially as a couple of months has passed since she died. I had hoped that time would settle things down (it has to an extent) and now it seems too late to seek help. No one offered / directed me to any support either so I wondered if not actually available?* (Bereaved daughter, RID024)
>
> *I am reluctant to reach out to bereavement services because I feel uncomfortable about the idea of making myself vulnerable to a complete stranger*. (Bereaved grandson, RID071)

In relation to GPs, people described difficulties getting appointments or feeling inadequately supported during telephone appointments. GP support commonly involved providing medication, helpline information and sick-notes for time-off work, with a lack of information about, or referral to, bereavement and mental health services also noted.

> *I am having difficulties coming to terms with my loss. GP can only offer medication, or helpline advice which has not helped really. I need company really*. (Bereaved husband, RID671)
>
> *I’ve found because there is no continuity of care at my own GP surgery that I have to make my case and talk about difficult things on the phone is so hard, especially when you have to make the case to a receptionist to start with, just to get to talk to a doctor…. All those things, like asking for help and finding the right words of what you’re going through, are hard. It’s easier not to do anything and to stick to your friends and family*. (Bereaved wife, RID458)

### Accessing support from friends and family

A substantial proportion of participants (n=279, 39%) reported difficulties getting support from friends and family. Across the sample, 25% (n=175) reported that their friends or family were unable to support them in the way that they wanted, whilst 19% (n=195) reported feeling uncomfortable asking for help. Three main themes were identified in free-text data: difficulties connecting and communicating with friends and family, disrupted grieving, and lack of understanding and empathy. These themes were again evident across gender and ethnic groups with other group-specific findings described.

#### Difficulties connecting and communicating with friends and family

Opportunities for in-person contact were minimised due to social distancing measures and geography. Needing physical comfort and ‘hugs’ was widely reported, with many describing difficulties talking openly about their feelings with friends and family, especially over the phone or internet. Isolation during lockdown and early bereavement made it harder to reconnect with friends over time. In some cases, isolation and disconnectedness were worsened by pre-existing strained relationships, or conflict surrounding end-of-life or early bereavement experiences.

> *I have not really sought support from family as they are affected too. We talk about Dad in a positive way, and joke about him as well. This helps. I would not “seek support” from any friends - what would I ask them? No idea. I suppose what could happen would be getting a bit drunk together and getting a few things off my chest, but this isn’t likely to happen in times of Covid*. (Bereaved son, RID340)
>
> *It’s been difficult for myself and my husband to get back into work and the pandemic related social isolation made it difficult for us to grieve openly with friends and family. This in itself put an emotional barrier up between us and them. As they hadn’t been through the unique trauma we’d experienced, it was difficult to set up a line of communication and build relationships again*. (Bereaved mother, RID267)

People worried about being a burden, and felt unable to trouble others also grieving. The perceived need to keep grief to one’s self and stay strong was particularly acute for parents, especially those home-schooling children during lockdown. Owing to the widespread stress caused by the pandemic, many feared adding to the emotional and mental health burden of others. People who lost elderly parents to non-COVID illnesses described further inhibitions about asking for help.

> *I’ve had to contend with managing my own grief with also supporting my children through theirs and dealing with a heavy workload, home-schooling and being unable to meet with my own friendship group or family for the support I would normally have looked for from them*. (Bereaved granddaughter, RID348)
>
> *There is no doubt that the COVID restrictions have made this period even worse than they would have been in “normal” times. It has sometimes made it hard to ask for help when I am aware that everyone is having a difficult time due to COVID and equally there are things that I might have been able to do to help myself - e*.*g. volunteering activities, which have not been possible. Two terrible life experiences happening at exactly the same time has been very hard and continues to be*. (Bereaved wife, RID469)

#### Disrupted grieving

Being unable to meet close family/friends for support disrupted the grieving process, making the death seem ‘surreal’ and harder to accept. A bereaved daughter described this as ‘a constant prolonging of a goodbye’ (RID112). Some felt that grieving families should have been permitted to meet during lockdown.

> *I think working through the anger and sadness I feel about how mum died and what we have consequently suffered in terms of loss of normal grief “rights”. (For example I have still not hugged my Dad or sister) is something that I need more help with now and I think the government has failed to take account of the damage done to bereaved families by not making allowances for them to have bereavement contact during lockdowns, crying on Zoom is just not the same. All I wanted when Mum first died was to get out of this house and go and have a cup of tea with a friend. I have been stuck in the house most of this year and for a long time with my husband and kids, home-schooling. I had no escape and nowhere to go*. (Bereaved daughter, RID734)
>
> *Just not being able to hug and be in the same room. After funeral I would have liked to have been in a room with my mother’s friends, my friends and family, sharing memories and stories, crying and laughing, etc. For a while I thought I would still organise a wake after COVID but now I think the moment has passed and that ritual will be missing too*. (Bereaved son, RID723).

Disruption to mourning rituals and collective support appeared especially salient for people from minority ethnic backgrounds, affecting extended family and community as well as immediate family.

> *Only my cousin’s wife [was] allowed to say good bye to her husband. His two children or any other relatives were not allowed. She returned home with her children. There was no other relatives there to support her (due to isolation and Covid regulations) [even] her mother was not there to [console] her. This was like pouring salt on wounds. Sharing ones grief reduces pain and help overcome the pain. Normally hundreds of relative would have been visiting her and sharing her grief which would have helped her and all of us to accept the death*. *But not being to visit her personally made it very difficult to overcome*. (Bereaved male cousin, RID653)
>
> *We have been unable to assist our relatives, especially us from Black Minority Ethnic (BME), in our background culture staying [close] with friends, visiting them frequently in time of bereavement is the most important thing we do. But right now, no one was [able] to do that which make the challenges even harder*. (Bereaved niece, RID680)

#### Lack of understanding and empathy

A lack of understanding and empathy within social networks was commonly described. Participants perceived friends and family members as feeling awkward and uncomfortable talking about grief or the deceased person, changing the subject or implying that they should have ‘moved on’. Many described receiving frequent calls in the first weeks of bereavement, but noted the decline as the months went on. One younger participant reflected that since parental loss was unusual in her peer group, her friends were unable to provide the support that she needed.

> *[Not] there in person, feel lonely while others get on with their lives where one doesn’t want to intrude. Also nobody who’s not lost a partner, can really understand…*.*It’s about time you pulled yourself together and got on with your life. But my life is gone, from language, to food, to walks in woods, to friendship and sharing*… (Bereaved wife/partner, RID111)

People also described how the exceptional nature of COVID-19 bereavement (including the anger associated with it) made it impossible for friends to understand, compounding their sense of loneliness and isolation. Experiences of social contacts disregarding regulations, questioning the seriousness of the pandemic or sharing conspiracy theories on social media were also distressing, further contributing to their alienation.

> *The covid bereavement group on Facebook have been a great source of comfort, as have immediate family. But whilst friends try to be helpful and kind - they don’t understand the anger which is also part of this grief. My Mother passed away 9 years ago from a severe stroke and whilst this was as big a shock as my father’s death - there was more of an acceptance about it. Friends offered tea and sympathy and a shoulder to cry on but then you picked yourself up and got on with it. With Covid it’s very different, the social isolation obviously doesn’t help but there is this underlying anger that not enough infection control procedures were put in place within our hospitals and therefore this death was avoidable!* (Bereaved daughter, RID635)
>
> *Other people don’t want to keep hearing it and some people who believe Covid is a hoax or conspiracy, it’s heart-breaking to have to listen to that crap continuously. People look at you like you are lying if you say Mum died if Covid. The ignorance out there is stifling sometimes*. (Bereaved daughter, RID318)

## Discussion

### Main findings

This study quantifies and describes the support needs and difficulties accessing support experienced by bereaved people during the COVID-19 pandemic. We identified high level needs for emotional and therapeutic support, alongside difficulties accessing both formal and informal support. Barriers included feeling uncomfortable asking for help and not knowing how to get help, as well as a lack of availability of support from bereavement services and GPs. Pandemic-specific challenges included high proportions of people perceiving social support to be inadequate, reduced in-person contact affecting the perceived quality and functionality of support, and disruption to collective mourning and grieving. The unique nature of pandemic bereavement and wider societal strains compounded the difficulties and isolation experienced by people bereaved during these exceptional times.

### What this paper adds

50 to 60% of bereaved people reported high or fairly high needs for help with processing feelings surrounding the death and loss, anxiety and depression, and communicating and connecting with friends and family, suggesting considerable needs for social/emotional support as well as reflective grief-focused support. Only a third perceived no need for bereavement service support due to adequate support from friends and family. This is significantly less than the 60% estimation in the pre-pandemic public health model(17,18), but validates recent pandemic-based predictions(13).

As in pre-pandemic studies, people commonly reported lack of understanding and compassion amongst friends and family, alongside difficulties expressing their feelings and asking for help(20-24). We found that these experiences have been exacerbated by the physical isolation and diminished opportunities for in-person support(25,26), the disruption to collective mourning caused by pandemic restrictions, as well as additional concerns over burdening others also experiencing hardship. Loneliness was compounded by the perceived uniqueness and anger associated with pandemic grief and COVID-19 deaths(36), alongside the distressing effects of people questioning or disregarding the pandemic.

However, whilst most participants felt that they needed additional support, most had not tried to access help from bereavement services or their GP. Strikingly, around three quarters of people with high/severe vulnerability were not accessing tier 2/3 support. As in pre-pandemic studies, reasons included lack of knowledge/information on how to get support as well as psycho-social barriers such as feeling uncomfortable asking for help(16,27). Some people felt less entitled to support at this time of crisis; others questioned the efficacy or appropriateness of the support on offer. Lack of face-to-face support and to a lesser extent COVID-specific support dissuaded people from taking up formal support. We identified preferences for support groups based on shared experiences or characteristics, the benefits of which have been described previously(16,19). For the 40% of people who did try to access bereavement service support, just over half experienced difficulties such as long waiting lists or ineligibility, supporting the findings of other studies(27-29). The main problems affecting the accessibility and quality of GP support were difficulties getting appointments or unsatisfactory telephone appointments(3,37), with inadequate signposting and referral to bereavement and mental health services also noted.

### Strengths and weaknesses

This mixed-methods study will be the first to longitudinally investigate peoples’ experiences of bereavement support during the Covid-19 pandemic in the UK. The baseline quantitative data demonstrates the extent of the difficulties experienced by the bereaved, whilst explanatory qualitative data provide rich insights into participant experiences. However, lack of random sampling means that the survey is not representative and by recruiting mostly online we were less likely to reach the very old or other digitally marginalised groups. Despite significant efforts and targeted recruitment, people from minority ethnic backgrounds and men are underrepresented in the data. Despite this, group sizes were sufficient to enable comparisons (although not to the level of specific ethnic groups), with group-specific observations reported where relevant in our qualitative findings. Quantitative analysis of the effects of demographic and clinical characteristics on support use is forthcoming.

### Implications for research

This study includes follow-up surveys at c.7 and 13 months post-death, longitudinal qualitative interviews and research exploring the impact of the pandemic on voluntary sector bereavement services and their response. Although our interviews target underrepresented participants such as those from minority ethnic backgrounds, same-sex couples and men, further research with these groups is needed. Research exploring the needs of bereaved children and young people, longer-term bereavement experiences, and experiences of statutory sector bereavement support providers is also recommended.

### Conclusions and implications for policy and practice

These results demonstrate high levels of need for emotional and therapeutic support, and the significant difficulties bereaved people face getting these needs met. Our results elaborate upon pre-pandemic inadequacies in formal and informal support, while demonstrating new pandemic-specific challenges including more complex, crisis-specific needs, diminished opportunities for face-to-face and group support, acute social isolation and disruption to collective grieving, and the wider societal consequences of the pandemic. Based on study findings we make three recommendations for improving the support available for bereaved people:

1. Increased provision and tailoring of services, including greater resourcing and expansion of national support as well as regional services in areas with long waiting lists. Safe ways to access face-to-face individual and group support as well as online and telephone support should be identified, with specific support available for groups with shared experiences and characteristics. This should include support which is culturally competent(16,30,31) and crisis/context competent(3,16). Training in core competencies specific to COVID-19 and identifying and sharing best practice amongst bereavement and palliative care providers would facilitate this.
2. Strategies to improve awareness of bereavement support options, including providing information on grief and bereavement services proactively following a death and ensuring accessible public information is available online and in community settings. GPs and other primary care providers should be better resourced to signpost bereaved patients to appropriate support(38).
3. More help with loneliness and isolation, including flexible support bubble arrangements for the recently bereaved when restrictions are in place(26). Following compassionate communities approaches(39,40), informal community-based interventions should be expanded, whilst educational and society level initiatives are needed to improve how, as a society, we communicate and support people experiencing death, dying and bereavement(40-42)

## Supporting information

CHERRIES checklist

## Data Availability

Full study data sets will be made available following study closure in February 2022. Data sharing requests will be considered prior to this and should be directed to Dr Emily Harrop

## Acknowledgements

Our thanks to everyone who completed the survey for sharing their experiences, and to all the individuals and organisations that helped disseminate the survey. We would also like to thank the project assistants, collaborators and advisory group members who are not co-authors on this publication: Dr. Emma Carduff, Dr Daniella Holland-Hart, Dr. Catriona Mayland, Prof. Bridget Johnston, Dr. Donna Wakefield

## Author contributions

E.H. and L.E.S. designed the study, led the application for funding and are co-principal investigators; E.H. drafted the paper; M.L., A.B., D.F., A.N., A.T.B., L.M., A.F., K.S., K.V.S., A.R., A.P., S.S. are members of the research team or the study advisory group and contributed to the design of the study and survey. D.F conducted the statistical analyses with data management assistance from M.L. and S.G., E.H., K.B., E.S. and S.G. conducted the thematic analysis of qualitative data. All authors contributed to drafting the paper and read and approved the final manuscript.

## Ethical approval

The study protocol and supporting documentation was approved by Cardiff University School of Medicine Research Ethics Committee (SMREC 20/59). The study was conducted in accordance with the Declaration of Helsinki and all respondents provided informed consent.

