## Supplemental File 2 for "Support needs and barriers to accessing support: Baseline results of a mixed-methods national survey of people bereaved during the COVID-19 pandemic"

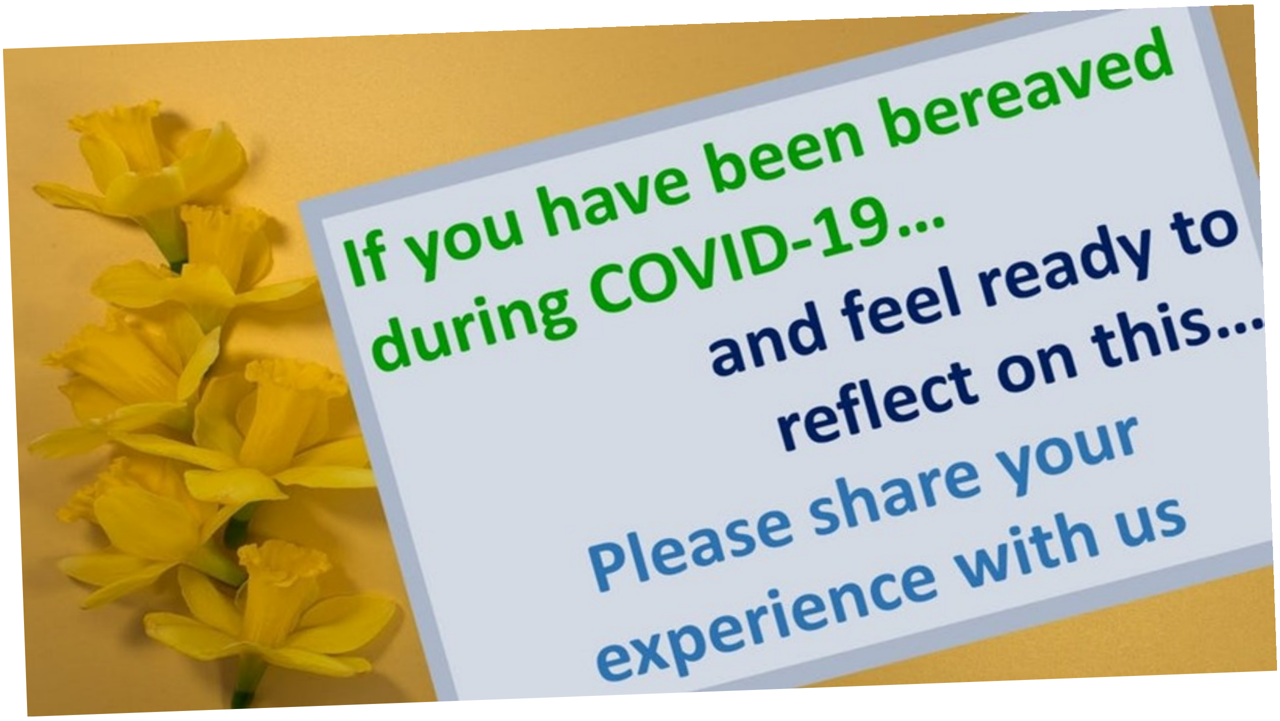


Cardiff University and the University of Bristol are conducting a **survey** looking at the **grief experiences** and **support needs** of people **bereaved** during the **pandemic.**

By conducting this survey we hope to identify ways of improving the care provided at the end of life and during bereavement.

If you have lost a loved one to COVID-19 or another cause of death during the pandemic, and would like to share your experience in our survey, please **click here** or visit www.covidbereavement.com

Or contact Emily Harrop:, **tel: 02920 687184** for further information or a paper copy of the survey.

### Thank you!

Cardiff University / Prifysgol Caerdydd
E·S·R·C Economic & Social Research Council
University of Bristol
Marie Curie — Care and support through terminal illness
Canolfan Ymchwil Gofal Lliniarol Marie Curie
Palliative Care Research Centre
CARDIFF CAERDYDD
